# Fully Homomorphic Collaborative Learning for Safe Cross-Healthcare Institution Development and Implementation of Foundation Models

**DOI:** 10.64898/2026.05.15.26353345

**Authors:** Song Bian, Hui Qiao, Tao Yan, Ziyao Xia, Xinyan Gao, Yuxin Xu, Ruiyu Shen, Taizhangtian Ma, Zhenyu Guan, Ya Xing Wang, Tien Yin Wong, Qionghai Dai

**Affiliations:** Department of Automation, Tsinghua University, Beijing, 100084, China; School of Cyber Science and Technology, Beihang University, Beijing, 100191, China; Institute for Brain and Cognitive Sciences, Tsinghua University, Beijing, 100084, China; Beijing National Research Center for Information Science and Technology, Tsinghua University, Beijing, 100084, China; School of Clinical Medicine, Tsinghua Medicine, Tsinghua University, Beijing, 100084, China; Beijing Visual Science and Translational Eye Research Institute (BERI), Beijing Tsinghua Changgung Hospital, Tsinghua Medicine, Tsinghua University, Beijing, 100084, China; Beijing Tsinghua Changgung Hospital Eye Center, Tsinghua Medicine,Tsinghua University, Beijing, 102218, China; Beijing Key Laboratory of Intelligent Diagnostic Technology and Devices for Major Blinding Eye Diseases, Tsinghua Medicine, Tsinghua University, Beijing, 100084, China; Singapore Eye Research Institute, Singapore National Eye Centre, 168751, Singapore

## Abstract

Foundation models (FMs) are powerful tools to allow the broad clinical application of artificial intelligence (AI) in healthcare systems, offering adaptability to different disease, modalities and clinical settings. However, FMs require large-scale datasets to train and fine-tune, while most real-world data are localized in siloed healthcare settings with strict data privacy protection, a restriction that poses a fundamental challenge in the cross-healthcare institution development of FMs. Here, we develop a fully homomorphic collaborative learning framework, named as FOCAL, that enables secure FM-driven diagnosis without exposing raw patient information. Different from traditional federated learning (FL) frameworks that aggregate locally trained models, FOCAL integrates fully homomorphic encryption (FHE) with split training to effectively execute collaborative learning completely over encrypted data. Specifically, we apply FOCAL on different types of retinal and pathology FMs to demonstrate its clinical performance. When facing gradient inversion attacks, FOCAL reduced the data leakage rate from 90.6% to 0% with comparable accuracy performance of the state-of-the-art FL paradigms, owing to the provable security provided by FHE. Moreover, under the same level of security, FOCAL can boost the macro-average AUROC by nearly 50% (from 0.5202 to 0.9831) when evaluated against fully encrypted FL models. In the multi-institution comparative experiments, FOCAL consistently outperforms all single-institution FMs, improving AUROCs by 9.62% and 14.46% on the ocular disease diagnosis and severity classification, respectively. Lastly, external validations on both retinal and pathology FMs further verified the accuracy and security advantages of FOCAL and highlighted its reliable interpretability and generalizability for cross-institution clinical development and implementation of FMs. FOCAL is a novel method to build a secure data-sharing AI community, facilitating healthcare institutions to benefit from and contribute to next-generation FMs development without compromising patient privacy and data security.

## Introduction

The application of artificial intelligence (AI) in the domain of medicine is one of the main driving forces to revolutionize modern healthcare systems. To achieve consistent performance across diverse downstream clinical tasks, medical AI is rapidly evolving from small, disease-specific and task-specific models to large foundation models (FMs)^[1–6]^ in fields such as radiology, ophthalmology^[7–9]^, pathology, dermatology, and other disciplines. However, clinical deployment of such FMs is dependent on broad generalizability across patient population and healthcare settings, which requires large-scale, multi-institutional, multi-ethnic patient population datasets for pretraining^[10,11]^. Unfortunately, medical data collection is not only costly to collect, but also heavily regulated^[12–14]^. Privacy laws, geopolitical tensions, and country-specific regulatory regimes create serious barriers to aggregating large-scale, diverse medical datasets. Therefore, the secure sharing of data across clinical sites has become an essential challenge for the next stage of medical AI development and adoption^[15–19]^.

The current method of federated learning (FL) is a promising solution to overcome data security issues across hospitals, research centers, and geographic regions by enabling collaborative model training and inference without centralized data aggregation^[20–22]^. Through FL, institutions can jointly learn generalizable medical representations from clinical data distributed across different data institutions. Nevertheless, as shown in Figure 1a, traditional FL still suffers from a number of critical limitations, such as reduced usability caused by heterogeneous hardware resources across clinical sites, degraded accuracy performance due to non-independent and identically distributed datasets, and security vulnerabilities against data-leakage attacks^[23]^. In particular, from the shared gradients or model parameters in traditional FL, malicious eavesdroppers can reconstruct sensitive patient information, such as medical images or text records^[24–26]^, posing significant risks to data confidentiality and regulatory compliance. In addition, distributed model training imposes substantial computational and resource burdens on each of the participating hospitals and medical institutions^[27]^. Thus, FL has not been a scalable approach for development of FM and other AI systems across global healthcare networks.

**Figure 1.**
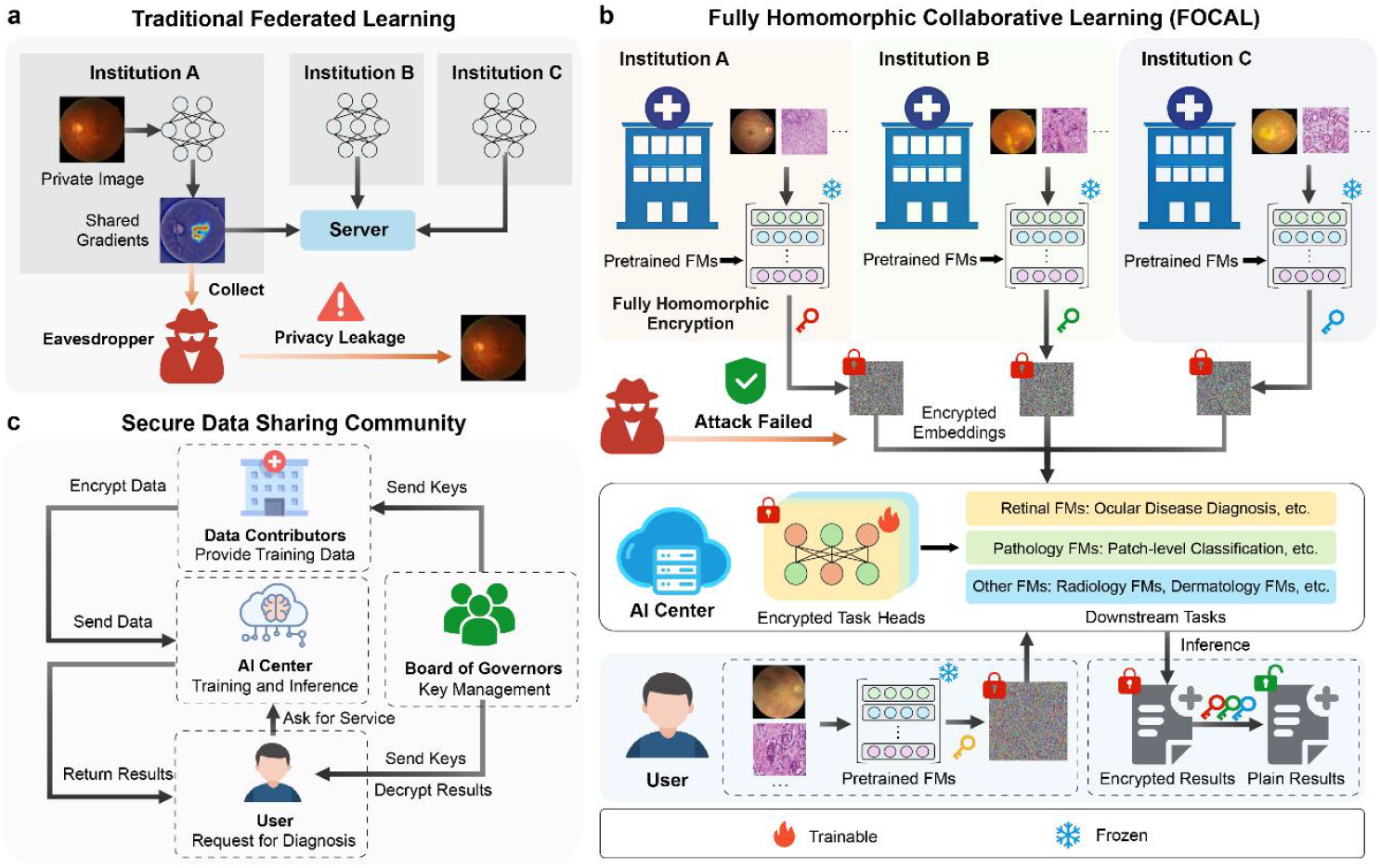
Overview of the FOCAL framework. **a**, Traditional FL exposes gradient information to the aggregating server, which allows a malicious server or third-party adversary to reconstruct private images from shared gradients. **b**, The proposed FOCAL framework that enables collaborative model fine-tuning and inference while keeping all images encrypted. During training, the data-contributing institutions holding private images first produce the feature embeddings using a frozen pretrained FM. Then, the embeddings are encrypted locally and then sent to the AI center. The AI center performs training of the encrypted task head, producing encrypted gradients for model updates, all over ciphertexts without decryption. During inference, the user encrypts the input images and send the ciphertexts to the AI center for encrypted inference. The encrypted predictions are returned to the user and decrypted them locally. Throughout the workflow, neither the data-contributing institutions nor the AI center ever get access plaintext inputs or feature embeddings, ensuring provably-secure protection of both institutional data and user privacy. **c**, For large-scale adaptation, FOCAL employs a four-party protocol for the construction of a secure data-sharing community. First, data contributors provide the encrypted training data and labels. Second, the AI center trains and serves FHE-protected models. Third, the users request encrypted diagnostic services. Lastly, a board of governors are responsible for key management and final data decryption for the authorized users.

Here, we propose a novel approach, FOCAL (fully homomorphic collaborative learning), a provably secure AI framework designed to continuously evolve the capabilities of medical FMs over multi-institution private datasets. FOCAL integrates fully homomorphic encryption (FHE)^[28,29]^ with split learning^[30]^ to allow encrypted models to be trained within a centralized AI hub without involving any plaintext data. Using FHE, the training and inference of FMs can be carried out completely over ciphertext without decryption, where the security of such ciphertexts is provable through mathematical reductions. Thus, data contributors and users of FOCAL only need to provide encrypted embeddings, thereby eliminating heavy local computation and training overheads. Furthermore, a jointly governed board of model developers oversees the cryptographic key management process to ensure transparency, safety, and fairness.

To test the effectiveness of this approach, we validate FOCAL on retinal and pathology FMs for the diagnosis for different types of diseases. We showed the following. First, we demonstrated that the proposed framework effectively safeguards sensitive patient information against data-leakage attacks during collaborative training and inference. When applied to 500 color fundus photos (CFPs), the gradient inversion attack can recover 90.6% of the input CFPs with structural similarity indices (SSIM) larger than 0.5, indicating a high data leakage rate. In contrast, attacks on the ciphertext embeddings produced by FOCAL yielded only noise-like outputs, with no reconstructed photos achieving any meaningful recovery of the private input CFPs. Second, we validated the advantages of FOCAL in safely leveraging multi-institution data on two ocular disease diagnosis tasks and one pathological classification task. Compared with encrypted FL, FOCAL shows improvements in macro-average AUROC by nearly 50% (the macro-average AURCO improved by 0.4629), while achieving a 192-fold reduction in model size and 168-fold faster inference speed for ocular disease diagnosis. Compared with locally-trained model, FOCAL consistently improves the AUROC by at least 9.62% for the disease diagnosis task and 14.46% for the disease severity classification task. Finally, on the external validation datasets, FOCAL maintains the accuracy advantages on both ocular and pathological diagnosis tasks, confirming its generalizability and robustness. Overall, FOCAL establishes a new paradigm for data secured, data efficient, and collaborative medical AI training and deployment. By enabling privacy-preserving knowledge sharing among distributed healthcare institutions, FOCAL lays out the foundation for a shared, AI-connected medical ecosystem, making global healthcare more accessible, affordable, and secure.

## Results

### Overview of the FOCAL Framework

As outlined in Figure 1b, the FOCAL framework involves three main participating parties: the medical institutions who hold private training data, the user who wish to perform inference on the trained models, and the AI center who serves as the computational platform to both the user and the medical institutions. Before training, a public pretrained FM is first distributed as an image encoder to each of the medical institutions. Next, each institution encodes their private data to produce feature embeddings using the same distributed FMs. Then, the embeddings and the associated data label are encrypted through FHE to produce the ciphertext inputs to the AI center for encrypted training. Upon gathering the encrypted embeddings and labels, the AI center performs the model training process entirely over FHE ciphertexts without decryption. The entire protocol is shown in Extended Data Figure 1.

As summarized in Table 1, existing privacy-enhancing technologies (PETs) that are commonly used in collaborative learning can roughly be classified into three main categories depending on their roots of trust: noise-based, hardware-based, and cryptography-based. Noise-based PETs, such as differential privacy^[31]^ and noise-based image encryption^[32,33]^ protect the privacy of the input data by adding randomized noises to disturb the data distribution. While noise-based PETs enjoy decent utility and low latency overheads, the security guarantees for most of the noise-based PETs rely on statistical obfuscations, which are not provably secure by the cryptographic standards. For example, in the recoverable image encryption scheme^[32]^, highly recognizable images can be recovered directly from encrypted data using a chosen-plaintext attack, which violates the basic requirement of semantic security for data encryption^[34,35]^. Similarly, hardware-based PETs, such as trusted execution environments^[36]^, leverage trusted hardware enclaves to preserve data security during model training and inference. However, since the encrypted data are in fact processed in a decrypted form inside the trusted hardware enclave, such methods are generally not considered compliant to the data security regulations such as the general data protection regulation (GDPR)^[13]^. Lastly, cryptography-based PETs provide provable security to data confidentiality, which is particularly useful in ensuring legal compliance for inter-regional collaborative learning scenarios. Within the cryptography-based PETs, FHE permits arbitrary computation to be carried out over ciphertexts and is known to have much lower communication overheads when compared to the secure multi-party computing techniques. Thus, by simultaneously achieving provable security and high usability, FHE is chosen as the cryptographic foundation for establishing FOCAL, where large numbers of medical institutions and service users can simply interact with a centralized AI service provider, i.e., the AI center, to jointly execute the training and inference protocols.

**Table 1.** Privacy-enhancing technologies used in collaborative learning.

| Technical Approach | Computation Speed | Communication Overhead | Security Basis |
| --- | --- | --- | --- |
| Differential Privacy | Extremely Fast | Low | Noise Statistical Property |
| Trusted Execution Environment (TEE) | Extremely Fast | Low | Hardware Root of Trust |
| Secure Multi-Party Computation (MPC) | Fast | High | Provable Security |
| Fully Homomorphic Encryption (FHE) | Slow | Low | Provable Security (Single Hardness Assumption) |

By offloading the computationally intensive training process to the AI center, FOCAL enables even resource-limited medical institutions to participate as data contributors and benefit from the shared outcomes of the framework. To support large-scale collaborative training, reduce the computational burden of the AI center, and mitigate noise accumulation in FHE ciphertexts, FOCAL applies low-rank adaptation (LoRA)^[37]^ to the encrypted task head to train only a small proportion of the weight parameters. Consequently, FOCAL can substantially lower the computational cost and participation barrier for potentially large numbers of participating institutions. To carry out the inference over FOCAL, the user performs similar encoding and encryption procedures to generate encrypted embeddings. The AI center can then carry out the inference over the encrypted task head using the input embeddings to homomorphically compute the inferenced label in ciphertext. Lastly, the user can decrypt the label ciphertext to obtain the inference result.

In real-world applications, FOCAL enables a large number of medical institutions and users to collaboratively train and inference on the encrypted task head. The security and fairness of large-scale collaborations need be properly managed. Therefore, as shown in Figure 1c, a board of governors (BOG) is formed within the participating parties to manage cryptographic keys and identities. First, the BOG is responsible for distributing the encryption keys in advance to the medical institutions (i.e., the data contributors) and the users before the training and inference processes. After a user performs a round of inference on the encrypted model, the BOG also needs to verify the identity of the user to permit a decryption on the results, a measure designed to prevent security risks arising from a single user managing the global decryption keys. By having a BOG to supervise all the participating parties in the FOCAL protocol, we essentially establish a new type of knowledge-sharing community to safely democratize and implement AI-based diagnosis services in clinical practices. A concrete characterization of the FOCAL protocol is provided in the Method section.

### Performance Validation on Ocular Disease Diagnosis Tasks

To validate the performance, we first applied FOCAL to a multicategory ocular disease diagnosis task based on the RETFound^[7]^ model.

#### Experiment Setup

We collected seventeen publicly available retinal image datasets along with three private clinical cohorts covering a diverse set of ophthalmic diseases and patient demographics across different geographic locations. As shown in Figure 2a and Extended Data Table 1, eighteen datasets were used for internal model fine-tuning, and the other two were held out as external validation datasets. We simulated a collaboration among three independent institutions: A, B and C, each containing data with varying disease types and patient population characteristics. Each of the three datasets contains data samples covering a variety of ocular diseases, including diabetic retinopathy (DR), glaucoma, age-related macular degeneration (AMD), cataract, retinal vein occlusion (RVO), and pathological myopia (PM). Together, these diseases encompass the primary causes of vision impairment and provide a representative assessment for the clinical applicability of a retinal FM.

**Figure 2.**
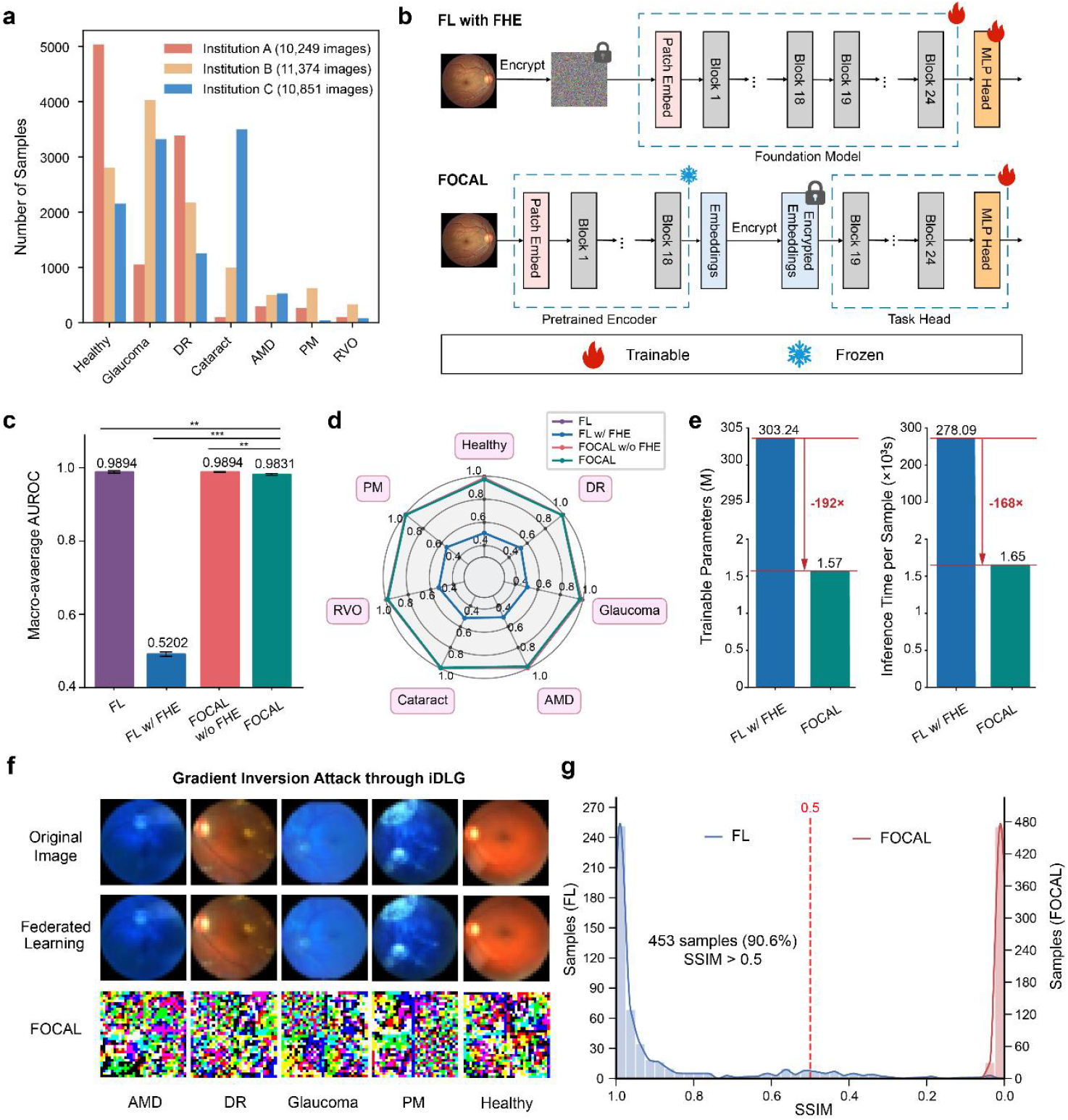
Comparative evaluation of FOCAL and FL. **a**, Dataset distribution across three institutions (A, B, C), covering seven classes (Healthy, DR, Glaucoma, AMD, Cataract, RVO, PM) with varying sample sizes per class. **b**, Workflow of FL with full-model fine-tuning and FOCAL with split fine-tuning under FHE. **c**, Macro-average AUROC comparison between FL and FOCAL with and without encryption. Without encryption, both methods achieve comparable performance. However, under FHE, FOCAL maintains high accuracy while FL experiences substantial degradation. Results were obtained by repeating each experiment with five random seeds. The bar center represents the mean value of AUROC and error bars represent 95% CI. Two-sided t-tests were conducted to compare FL and FOCAL under each condition, and significance levels were annotated in the plot using the convention: *** for *p* < 0.001, ** for *p* < 0.01, * for *p* < 0.05, and ns otherwise. **d**, Class-wise AUROC radar plots for FL and FOCAL. Across all seven classes, FOCAL sustains the performance advantage observed in subfigure c under encrypted settings. **e**, Efficiency comparison of FL and FOCAL under FHE. FOCAL reduces the number of trainable encrypted parameters by 192× and accelerates encrypted inference by 168× relative to full fine-tuning in FL. **f**, Privacy evaluation via the iDLG attack. In non-encrypted FL, private images can be reconstructed from shared gradients, while the encrypted FOCAL workflow prevents any meaningful recovery, thereby preserving data privacy. **g**, SSIM distribution of reconstructed images after 150-round iDLG attack. Notably, 90.6% of FL samples exceed 0.5 SSIM, indicating severe information leakage of the original input data image. Whereas, the SSIM for recovered images under FOCAL are well below 0.5, verifying its robustness against gradient inversion attacks.

While RETFound is typically adapted to downstream tasks through full-model fine-tuning, such approach becomes infeasible over private data that are distributed across multiple medical institutions. To solve the security challenge in an efficient manner, FOCAL applies split learning to update only a portion of the encrypted task head with LoRA, which minimizes computational overheads while preserving both data security and model performance. In Figure 2b, we provide a conceptual illustration for two types of encrypted training strategies on data from three institutions, namely, the FL model with fully encrypted fine-tuning (i.e., FL w/ FHE) and FOCAL. To enable provable security in traditional FL, the entire model parameters of the FM as well as the downstream task heads are required to be encrypted and uploaded to the AI center for fine tuning. In contrast, FOCAL splits the model into a public encoder (i.e., the FM) and an encrypted task head consisting of a considerably less amount of trainable parameters. Since the public encoder is frozen throughout the entire training and inference processes, the AI center only needs to fine-tune the lightweight encrypted task head. The detailed structure for the split FOCAL model is further explained in Extended Data Figure 2.

#### Comparing FOCAL with FL

As shown in Figure 2c and Figure 2d, when trained without FHE, the plaintext version of FOCAL achieves comparable performance to plaintext FL. However, when encrypted, we observed that FL with FHE experienced a sharp drop in average AUROC to 0.5202, while that of FOCAL retains at 0.9831. The main reason here is that, due to the large number of parameters that need to be trained in FL with FHE, a significant level of noises accumulates during the forward and backward propitiation processes, resulting in severely reduced accuracy performance for FL over FHE. In contrast, FOCAL only applies LoRA-based fine-tuning to the attention layers in the encrypted task head, which produced a negligible amount of computation noises. In addition, since the total amount of ciphertext computations are also reduced, we see that FOCAL can be up to 168× faster in inference time when compared to FL with FHE. The results indicate that split fine-tuning under FHE effectively preserves model utility while substantially reducing computational overhead, thereby providing a practical balance between data security and model usability.

To demonstrate the security properties of FOCAL, we use the improved deep leakage from gradients (iDLG) attack to examine whether gradient leakage can lead to breaches of the input datasets, where the attack strategy is detailed in Extended Data Figure 3. As illustrated in Figure 2f and Figure 2g, iDLG successfully reconstructed 90.6% recognizable CDFs from the gradients exchanged during the weight aggregation process of FL with SSIM larger than 0.5, confirming the privacy risks of conventional collaborative approaches. In contrast, when FHE is used, the encrypted gradients rendered inversion attacks ineffective, where all recovered images have SSIMs extremely close to 0. These results demonstrate that, by integrating FHE, FOCAL not only maintains competitive performance but also provides strong protection against gradient inversion attacks. This security advantage highlights the practical value of deploying FOCAL in real-world, privacy-sensitive multi-institutional collaborations.

#### Taking the Advantages of Secure Cross-Institution Learning

To demonstrate the effectiveness of cross-institutional collaborative learning, we compared two training paradigms in Figure 3: local full training of FM on a single-institution dataset in plaintext (i.e., Institution A or B or C) and encrypted split fine-tuning on the cross-institution dataset (i.e., FOCAL on A&B&C). Each single-institution model was trained and evaluated locally using only a single dataset of either A, B or C. Meanwhile, the FOCAL A&B&C model was trained using encrypted embeddings from all three datasets under the FOCAL framework. As shown in Figure 3a, when evaluated on single-institution datasets, both local training and FOCAL achieved comparable accuracy results. However, when evaluated on the combined cross-institution test dataset that contains test data from all three datasets, the FOCAL model exhibited a significant performance gain over the locally trained models, as shown in Figure 3b. In particular, the FOCAL on A&B&C model achieved an average AUROC of 0.9831 (95% CI 0.9807, 0.9855), substantially outperforming all single-institution fine-tuned baselines (0.7811–0.8968) with improvements of at least 9.62%. Such results demonstrate that, by adopting FOCAL, we can effectively mitigate domain bias by learning more generalizable representations across heterogeneous data sources by only training the downstream encrypted task head.

**Figure 3.**
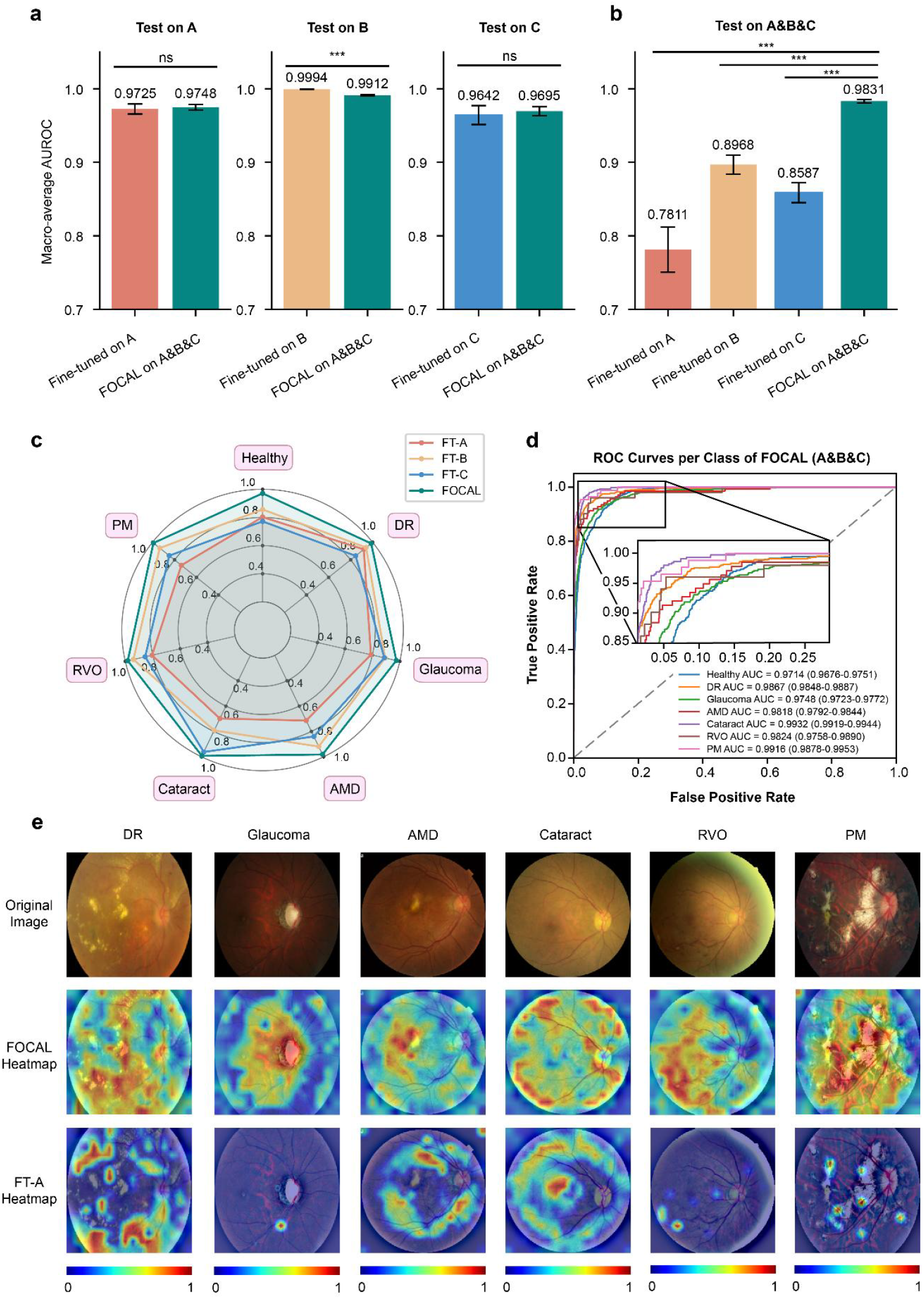
Comparative evaluation of single-institution plaintext full fine-tuning and the proposed FOCAL framework. **a**, Performance of locally trained single-institution models (FT-A, B, C) and the FOCAL model when evaluated on the test set in each of the institution’s own dataset. FOCAL achieves accuracy comparable to full plaintext fine-tuning performed independently at each institution, indicating that encrypted split fine-tuning does not compromise within-domain performance. The bar center represents the mean value of AUROC and error bars represent 95% CI. Two-sided t-tests were conducted to compare FL and FOCAL under each condition. Here, significance levels were annotated in the plot using the notation: *** for *p* < 0.001, ** for *p* < 0.01, * for *p* < 0.05, and ns otherwise. **b**, Evaluations on the aggregated cross-institution test set (A+B+C). The FOCAL model substantially outperforms all single-institution models, demonstrating its ability to leverage heterogeneous cross-institutional information and mitigate domain bias. Results were obtained by repeating each experiment with five random seeds. **c**, Class-wise comparisons on the seven-class disease diagnosis task tested on the aggregated dataset (A&B&C). Radar plots show that across all disease classes, FOCAL consistently surpasses single-institution models, reflecting stronger robustness to inter-domain variability. **d**, ROC curves of the FOCAL model across all disease types on the combined test set. High and stable AUROC values across classes further confirm the model’s reliable performance in multi-disease predictions. **e**, Score-CAM visualizations comparing single-institution models and the FOCAL model. Heatmaps show that FOCAL attends to clinically meaningful retinal regions and provides more consistent and diagnostically relevant attention patterns than single-institution models.

Figure 3c inspects the detailed performance comparisons of FOCAL against local training over the seven classification labels. For all disease categories, FOCAL consistently achieves higher macro-average AUROC values than all models locally trained on single-institution datasets. Notably, the largest gains are observed in categories with more diverse phenotypic presentations, such as DR and AMD, where single-institution training suffers from limited data variability. Additionally, the ROC curves in Figure 3d and Extended Data Figure 4 also show similar results, confirming the prediction stability of FOCAL across different ophthalmic disease types. In short, we see that, despite operating over an encrypted version of a split task head, FOCAL is able to consistently surpass the accuracy of plaintext full fine-tuning. More importantly, the above results highlight the importance of having a cross-institution collaborative learning framework that permits the share of clinical expertise across the medical community in a secure and performant manner.

To better comprehend the interpretability of FOCAL, we implemented the score-weighted class activation mapping (Score-CAM) technique on the split model to visualize the diagnostically critical regions that drive the decision-making process^[38]^. In our implementation, the target layer was the normalization layer of the last transformer block in ViT^[39]^ as shown in Extended Data Figure 5. As sketched in Figure 3e, By highlighting regions of CFPs that contributed most to the prediction results, the heatmaps produced by Score-CAM provided intuitive insights into whether the agent focuses on clinically relevant features. As illustrated in Figure 3e, by securely gathering cross-institutional data for model fine-tuning, the focus of FOCAL accurately covers the pathological region, and shows improvements to the single-institution case. More concretely, for DR, the heatmaps consistently emphasized microaneurysms, intraretinal hemorrhages, and exudates, aligning with established diagnostic criteria across different disease stages. In glaucoma cases, the model primarily attended the optic disc and cup regions, where structural changes such as increased cup-to-disc ratio are critical indicators. For AMD, the highlighted areas were concentrated around the macula, especially regions with choroidal neovascularizations and scars. In cataract classification, the attention diffused across the posterior pole, reflecting the blurred retinal details captured in images of affected patients. Note that, in the glaucoma, AMD and cataract experiments, the single-institution model FT-A produced incorrect prediction results, thereby showing poor interpretability. Similarly, retinal vein occlusion was characterized by heatmaps focusing on vascular abnormalities and intraretinal hemorrhages, while in pathological myopia, the model consistently concentrated on peripapillary atrophy and myopic maculopathy. The observations show that the FOCAL model not only achieves high AUROC performance but also grounds its predictions in pathologically meaningful image regions.

### External Validations on Disease Diagnosis Tasks

To further assess the generalizability of FOCAL in real-world deployment scenarios, we evaluated the locally-trained single-institution models (FT-A, FT-B, FT-C) and the FOCAL model on two external datasets, AOD^[40]^ and RFMID^[41]^ (Extended Data Table 1). These datasets differ substantially in scale and disease composition, providing complementary tests of robustness for the AI models. Specifically, AOD contains 13,081 color fundus photos across six categories (healthy, DR, glaucoma, AMD, cataract, PM), and is one of the most heterogeneous public datasets for ophthalmic disease diagnosis. RFMID, by contrast, includes 1,079 CFPs spanning four diagnostic categories (DR, AMD, RVO, PM), and is widely used as a benchmark for assessing model generalizability under limited-sample conditions.

We divide the external validation experiment into three sets of evaluations. First, as shown in Figure 4a, FOCAL achieved the highest macro-average AUROC on AOD, outperforming all single-institution models with clear statistical significance. Per-class ROC curves further demonstrate that FOCAL maintains strong discriminatory ability across all six disease categories. Although the AUROCs of FOCAL were slightly lower than FT-B on a small subset of categories, this discrepancy likely reflects the substantially larger domain diversity in AOD, where the local model trained on institution B coincidentally aligns more closely with the data distribution of the particular class. We note that, even in such cases, the performance of FOCAL remained comparable rather than degraded, reinforcing the benefit of aggregating heterogeneous multi-institutional knowledge. Second, FOCAL demonstrated stronger advantages over single-institution models on the RFMID dataset (Figure 4b). The model consistently surpassed all single-institution fine-tuned models across all four diseases, achieving the highest macro-average AUROC. The relatively small size of the RFMID dataset exacerbated overfitting in locally-trained models, whereas FOCAL retained stable decision boundaries that generalized effectively to unseen cohorts. Lastly, to examine the concrete decision boundaries, we sketched the t-SNE plots in Figure 4c to compare the feature embeddings of FOCAL against one of the single-institution models (i.e., FT-A). It is clear that FOCAL produces more compact and well-separated feature clusters across both AOD and RFMID datasets than single-institution models. Full visualizations across all models are provided in Extended Data Figure 6.

**Figure 4.**
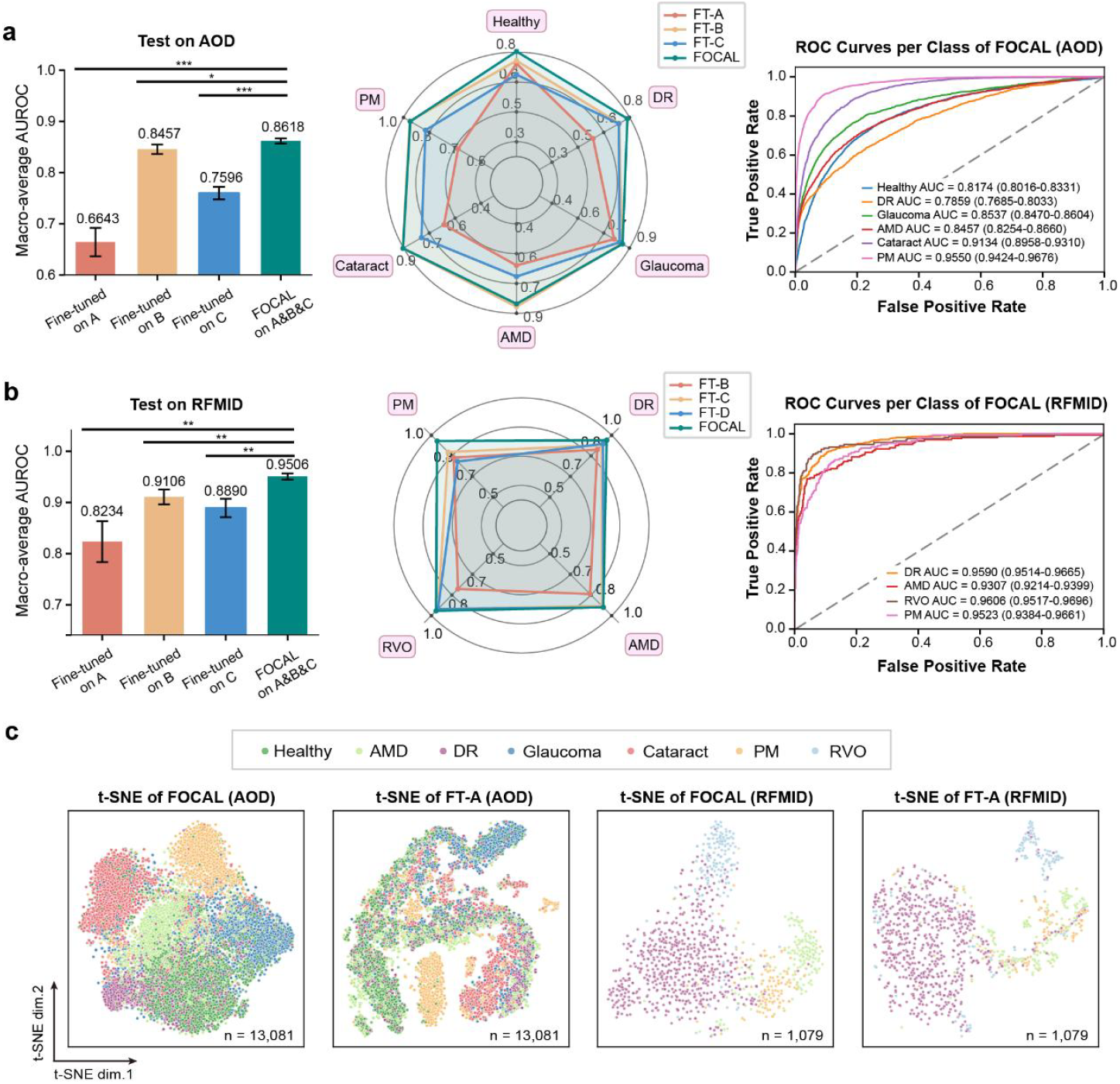
External validation of FOCAL on two real-world ophthalmic datasets. **a**, Performance comparison on the AOD dataset. FOCAL achieves the highest macro-average AUROC with statistically significant improvements over all locally fine-tuned models (FT-A, FT-B, FT-C). The radar chart illustrates that FOCAL maintains consistently superior or comparable performance even when single-institution models occasionally align better with specific domain distributions. Per-class ROC curves also show strong discriminative ability across all six disease categories. The bar center represents the mean value of AUROC and error bars represent 95% CI. Two-sided t-tests were conducted to compare FL and FOCAL under each condition, and significance levels were annotated in the plot using the convention: *** for *p* < 0.001, ** for *p* < 0.01, * for *p* < 0.05, and ns otherwise. **b**, External validations on the RFMID dataset. FOCAL outperforms all single-institution models across the four diseases and achieves the highest macro-average AUROC, demonstrating enhanced robustness under limited-sample and cross-domain conditions. **c**, t-SNE visualization of feature embeddings. Compared with a representative single-institution model (FT-A), FOCAL produces more compact and better-separated clusters across external datasets, indicating improved alignment between learned representations and underlying disease structure.

### External Validations on Severity Classification Tasks

In addition to the diagnosis tasks described above, we evaluated FOCAL on the DR severity classification task, which is a critical application of AI-based clinical diagnosis for the finer discrimination of pathological changes^[42–44]^. Accurate gradings of mild, moderate, severe, and proliferative are essential for the screening, referral prioritization and treatment planning of DR, rendering the task a representative benchmark for assessing general applicability of FOCAL for clinical practices. Here, we collected and grouped six public datasets into three simulated institutional cohorts (DR1, DR2 and DR3) for the training under the same two paradigms used in the disease diagnosis experiments: single-institution full fine-tuning over plaintext and cross-institution split fine-tuning with FOCAL over encrypted data. Two additional datasets, SYSU^[45]^ and DRA^[46]^, were held out for external validation, and the detailed dataset composition is explained in Extended Data Table 3.

Across the multi-institution test set formed by aggregating DR1, DR2 and DR3, FOCAL obtained the highest macro-average AUROC, surpassing all single-institution baselines (Figure 5b, left) with improvements of at least 14.46%. This performance advantage reflects the benefit of securely leveraging heterogeneous data from multiple institutions, which enables the model to learn more diverse disease manifestations than any single-institution dataset alone (detailed ROC curves are provided in Extended Data Figure 7). External validations using the SYSU and DRA datasets further confirmed the trend. On both datasets, FOCAL achieved higher macro-average AUROC than all locally trained models (Figure 5b, middle and right), demonstrating stronger robustness to distributional shifts. We make a special remark on the comparison on the DRA dataset here: although FOCAL did not outperform FT-A by a statistically significant margin, it still delivered the highest AUROC overall. This outcome is attributable to DRA’s substantially larger test size (9,316 images), which reduces performance variance and makes small inter-model differences statistically harder to distinguish. Even under such stricter evaluation setting, FOCAL maintained superior performance, highlighting its stability across large and heterogeneous external cohorts. Additionally, as shown in Figure 5d, we also verified the interpretability of FOCAL through generating heatmaps for each DR severity grade using external datasets (SYSU and DRA). For each severity level, an original image is paired with its corresponding Score-CAM heatmap that highlights the lesion areas microaneurysms, intraretinal hemmorrage, hard exudates, cotton-wool spots and epiretinal proliferative membranes) detected by the FOCAL model.

**Figure 5.**
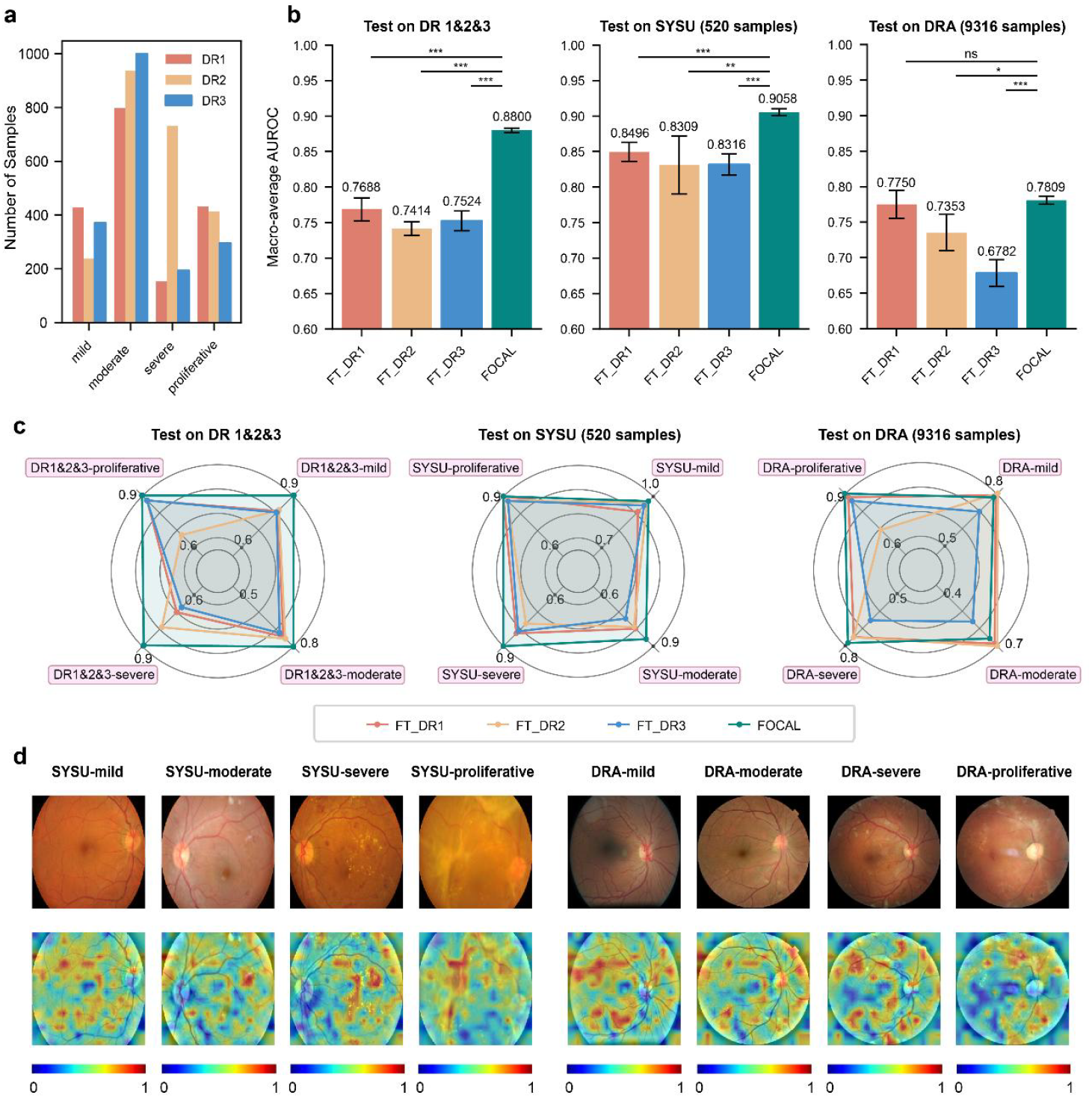
Diabetic retinopathy grading performance of FOCAL across multi-institution and external test sets. **a**, Datasets used for the DR grading task. Three virtual institutions (DR1–DR3) were constructed from six publicly available DR datasets, each providing heterogeneous imaging characteristics and severity distributions. **b**, Comparisons between single-institution models and the FOCAL model. On the aggregated multi-institution test set, FOCAL achieved the highest macro-average AUROC, substantially outperforming all single-institution baselines. External validations on SYSU and DRA show that FOCAL consistently outperformed all locally trained models, demonstrating enhanced robustness under distributional shift. Although the improvement over FT-A on DRA was not statistically significant due to the dataset’s large sample size, FOCAL still achieved the highest AUROC overall. Results were obtained by repeating each experiment over five random seeds. The bar center represents the mean value of AUROC and error bars represent 95% CI. Two-sided t-tests were conducted to compare FL and FOCAL under each condition, and significance levels were annotated in the plot using the convention: *** for *p* < 0.001, ** for *p* < 0.01, * for *p* < 0.05, and ns otherwise. **c**, Radar plots comparing FOCAL with single-institution fine-tuning. Across DR1 to DR3 on both external datasets, FOCAL achieved consistently higher AUROC for all severity levels, confirming the benefits of incorporating heterogeneous multi-institutional data. **d**, Score-CAM heatmaps of FOCAL on the two external datasets. FOCAL produces stable and clinically aligned attention patterns across diverse imaging conditions.

In conclusion, the results of the DR grading task align closely with the findings from the multi-disease diagnosis experiments: secure collaborative learning through FOCAL leads to consistently stronger performance and generalizability than models trained independently at single institutions.

### Performance Validation on Patch-level Pathological Image Classification Tasks

To further demonstrate the generalizability and robustness of FOCAL, we extended our validation to the computational pathology task which involves different medical imaging modalities. Specifically, we employed UNI^[47]^, a state-of-the-art general-purpose FM for pathology, to perform a patch-level esophageal carcinoma (ESCA) tissue classification task, as illustrated in Figure 6a.

**Figure 6.**
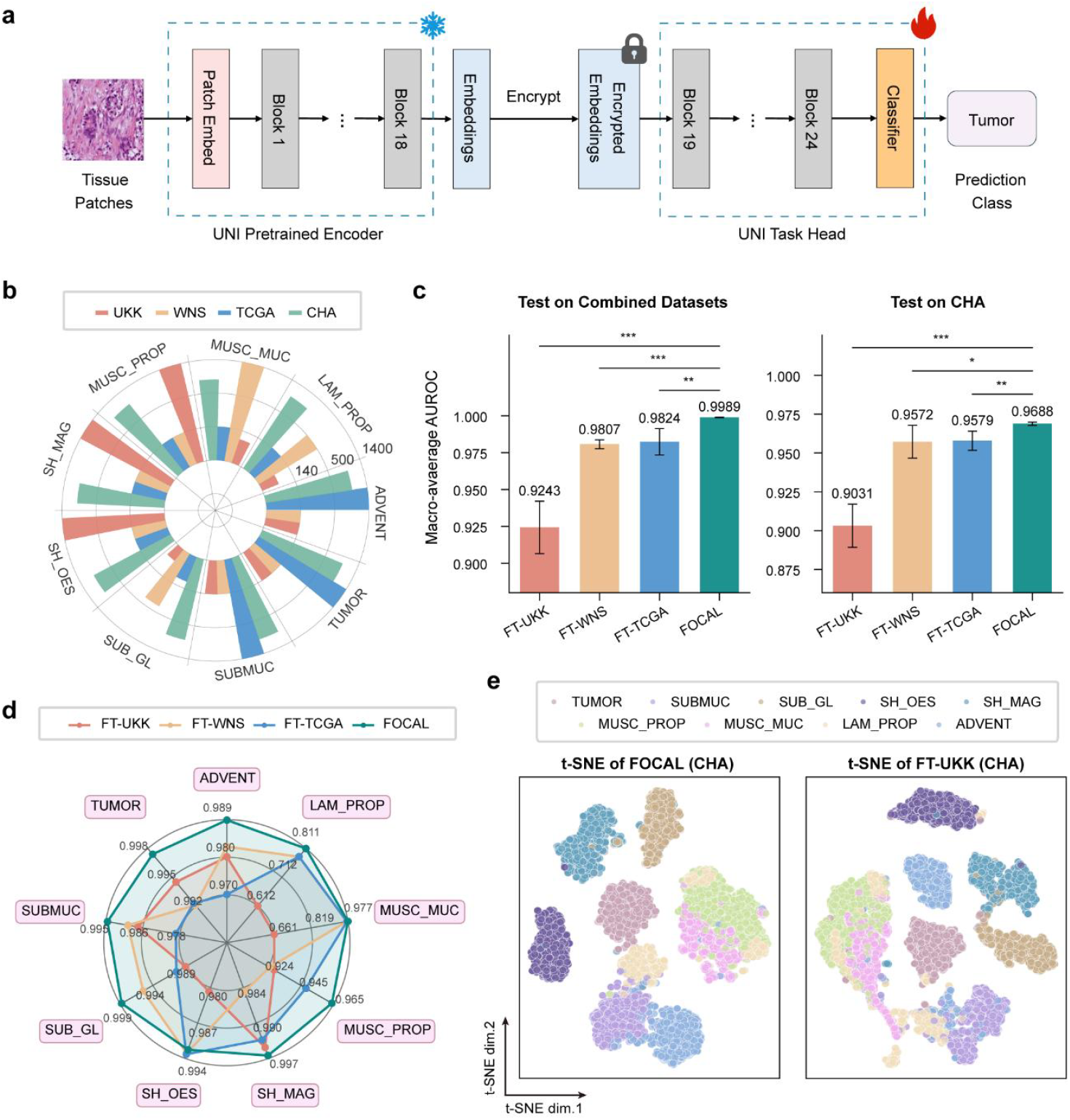
ESCA tissue classification performance of FOCAL across multi-institution and external test sets. **a**, Illustration of the patch-level ESCA tissue classification task utilizing the UNI as the backbone in FOCAL. **b**, Data distribution of the nine tissue classes across the three data-contributing institutions (UKK, WNS, and TCGA) and the external institution (CHA) in our experiments. **c**, Comparison of macro-average AUROCs between FOCAL and single-institution models. The left panel displays performance on the combined internal test set (UKK + WNS + TCGA), while the right panel demonstrates generalization capability on the unseen external CHA test set. The bar center represents the mean value of AUROC and error bars represent 95% CI. Two-sided t-tests were conducted to compare FL and FOCAL under each condition, and significance levels were annotated in the plot using the convention: *** for *p* < 0.001, ** for *p* < 0.01, * for *p* < 0.05, and ns otherwise. **d**, Radar plot illustrating class-wise AUROC performance on the external CHA dataset. FOCAL consistently achieves better performance across eight tissue categories compared to models trained on individual datasets. **e**, t-SNE visualization of feature embeddings on the external test set, comparing FOCAL-trained model with the model trained solely on the UKK institution.

We utilized a large-scale, multi-center ESCA dataset^[48]^ comprising histopathology regions of interest (ROIs) extracted from whole-slide images (WSIs). To simulate a realistic collaborative learning scenario characterized by data heterogeneity, we constructed a distributed environment involving three distinct data contributors: University Hospital Cologne (UKK), Landesklinikum Wiener Neustadt (WNS), and The Cancer Genome Atlas (TCGA). A fourth center, University Hospital Berlin – Charité (CHA), is used as an external testing cohort. We curated a subset of the dataset containing 9 representative tissue classes to create heterogeneous data distributions among the contributors. As shown in Figure 6b, the data distribution varies significantly across institutions, mimicking the real-world data island challenge (detailed dataset compositions are provided in Extended Data Table 5). Following the experiment settings in the ocular tasks, we compared FOCAL trained collaboratively over multi-institution datasets against models trained on single-institution datasets.

The evaluation results demonstrated the clear performance advantages of FOCAL in both internal and external testing environments. As shown in Figure 6c, FOCAL consistently achieved a significantly higher macro-average AUROC compared to any single-institution model. Notably, in the external validation on the CHA dataset, FOCAL effectively mitigated the overfitting issues common to single-center training, demonstrating generalizable performance. As shown in the radar chart of class-wise AUROCs on the external CHA test set (Figure 6d), FOCAL outperformed all single-institution models across eight tissue categories, confirming that its improvements are uniform and not driven by a specific subset of classes. In addition, t-SNE visualizations of feature embeddings on the CHA dataset (Figure 6e) reveal that FOCAL produces more compact and better-separated clusters than models trained on single-institution datasets (e.g., UKK), showing improved representation alignment and enhanced interpretability. Based on the above experiment results, we conclude that FOCAL and proves its capability to be applied to multiple types of FMs, unlocking the potentials of multi-center data in diverse clinical tasks, delivering substantial performance gain while being interpretable and privacy-preserving.

## Discussion

The implementation of AI-based medical diagnosis represents a crucial step toward achieving affordable and democratized healthcare. FMs offer a powerful base for AI-based healthcare systems as they can be adapted to multiple clinical tasks through fine-tuning. However, leveraging the full potential of FMs requires large-scale and diverse datasets that are often possessed by different medical institution, raising privacy and regulatory concerns over data sharing. To overcome utility-security dilemma, we introduced FOCAL, a framework that enables secure, privacy-preserving collaboration among healthcare providers to implement retinal FMs for clinical practices. By combining FHE with split learning, FOCAL enables FMs to collaboratively utilize private data across institutions without compromising confidentiality. Furthermore, the collaborative learning capability goes beyond a disease diagnosis application: we showed that the framework can easily be applied to both retinal and pathology FMs, Hence, allowing various types of medical FMs to continuously and securely learn from diverse and distributed medical datasets. Based on the proposed learning paradigm, we provide a scalable pathway for transforming isolated institutional resources into a collaborative and intelligent learning community, democratizing AI-based diagnosis capabilities across the entire healthcare network.

To examine the performance of FOCAL, we conducted extensive multi-institution experiments on both retinal and pathology FMs. For the ocular disease diagnosis tasks, we employed seventeen public and three private ocular disease datasets encompassing diverse populations and imaging conditions. In the ocular disease experiments, the data were grouped to simulate three independent institutions, each representing a distinct healthcare provider. The results demonstrated that collaborative learning under FOCAL consistently outperformed both traditional federated learning and single-institution local training, particularly in external validations where models trained on combined encrypted data exhibited superior generalization to unseen datasets. Moreover, FOCAL supports the joint adaptation of FMs to multiple ophthalmic diseases within one unified pipeline, facilitating comprehensive and privacy-preserving clinical deployment without redundant fine-tuning across individual tasks or institutions. To validate cross-domain generalizability, we also deployed FOCAL on a pathology FM in the patch-level pathological task. We show that FOCAL maintained the accuracy advantages, indicating its potential as a unified, privacy-preserving solution for multi-disciplinary medical AI collaboration.

In addition to the performance benefits, this study also closely inspected the fundamental security properties of collaborative medical learning. To the best of our knowledge, FOCAL represents the first framework to incorporate FHE throughout the entire training and inference stages for medical FMs, where all computations are carried out in the ciphertext domain. To examine the effectiveness of FHE, we studied how model gradients can be used to recover the private inputs from the training institutions. In particular, prior research have only showed the effectiveness of gradient attacks, such as iDLG, over traditional convolutional neural networks. How such attacks can be transferred to FMs based on the transformer architecture remained obscure. In this work, we showed that, without encryption, most of the input images can be reconstructed with high precision from the shared gradients. Conversely, with FHE, gradient inversion attacks failed completely, where the recovered images were indistinguishable from random noises. We also note that data-leakage attacks become even more pronounced for FMs over split learning, as the openly available encoder weights further lower the barrier for model inversion. These findings provide concrete proof for adopting FHE as a strong and practical solution against medical data leakage, which is critical in establishing trustworthy cross-institutional medical AI collaboration.

Finally, to interpret the exact model behaviors under the fully encrypted split learning setting, we provide various types of data visualizations to confirm that the performance improvements of FOCAL are attained in an interpretable fashion. First, we visualized the heatmaps on partial model gradients to show that, compared to models trained locally on a single institution, models trained under FOCAL pay more attention to the pathological areas. Meanwhile, in the external validation experiments, it was also demonstrated that the feature maps produced by FOCAL models are better clustered, leading to substantially enhanced decision boundaries. Therefore, we conclude that, by employing proper security measures, medical FMs can indeed benefit from cross-institution collaborative learning in a safe and explainable manner.

Despite the encouraging results, this study has several limitations that define directions for future research. The current implementation of FOCAL focuses on a single case study involving retinal FMs trained over CFPs. While this modality provides valuable information for many ocular diseases, incorporating additional imaging sources such as optical coherence tomography^[49]^, ultra-widefield^[50]^ retinal images along with structured clinical metadata can substantially enrich the learned representations and extend diagnostic coverage. Moreover, although the present framework can address the main types of ocular diseases, integrating FMs across different medical specialties, such as cardiology, neurology, and oncology, can enable the formation of a community of broader scope, where diverse institutions collaboratively enhance the generalizability of multimodal medical AI systems without compromising data privacy.

In summary, we proposed FOCAL, a privacy-preserving collaborative learning framework for FMs. Our experiments show that FOCAL enables accurate, efficient, and secure training and inference of FMs across multiple institutions while maintaining data confidentiality. This work demonstrates a scalable approach to build a safe shared medical ecosystem and lays the groundwork for future research on privacy-preserving AI in healthcare.

## Methods

### Datasets

We compiled a comprehensive retinal image dataset from seventeen public and three private clinical sources, covering six common ophthalmic conditions: diabetic retinopathy (DR, including four severity levels), glaucoma, age-related macular degeneration (AMD), cataract, retinal vein occlusion (RVO), and pathological myopia (PM)^[40,41,45,46,51–64]^. In addition to these disease categories, normal CFPs were also included, resulting in a total of seven classification labels. Private datasets were collected from a specialized ophthalmic hospital, the ophthalmology department of a general hospital, and a medical examination center, representing a set of diverse clinical data sources.

To simulate real-world heterogeneity across healthcare providers, the combined dataset was split into three groups. During dataset splitting, we kept all data from each individual data source intact, i.e., we do not distribute samples from a single data source across different groups, thereby maintaining its intrinsic distribution and image characteristics. The grouping strategy ensured that each group contained representative cases of all six diseases as well as healthy samples, while maintaining comparable sample sizes. On average, each group comprised approximately 10,000 CFPs, which were then partitioned into training, validation, and test sets in a 7:2:1 ratio. Detailed dataset compositions and grouping strategies are explained in Extended Data Table 1.

For the diabetic retinopathy (DR) grading task, we selected six publicly available fundus datasets annotated with four DR severity levels (mild, moderate, severe, and proliferative). These datasets were grouped into three simulated institutional cohorts (DR1, DR2 and DR3), each containing approximately 1,800 CFPs to mimic multi-institution data heterogeneity. Within each cohort, the images were randomly partitioned into training, validation, and test subsets following a 7:2:1 ratio. In addition, two publicly available DR datasets (SYSU and DRA) were included solely for external validation to assess the generalizability of the proposed framework. Detailed dataset composition and per-cohort statistics are provided in Extended Data Table 3.

For the esophageal carcinoma (ESCA) tissue classification task, we utilized a multi-center dataset comprising histopathology ROIs annotated with nine representative tissue classes: ADVENT, LAM_PROP, MUSC_MUC, MUSC_PROP, SH_MAG, SH_OES, SUB_GL, SUBMUC, and TUMOR^[48]^. These samples were organized into three distinct data contributors to mimic realistic multi-institution data heterogeneity. Within each cohort, the ROIs were randomly partitioned into training, validation, and test subsets following a 7:1:2 ratio. In addition, the dataset from CHA was included solely for external validation to assess the generalizability of the proposed framework. Detailed dataset composition and per-cohort statistics are provided in Extended Data Table 5.

### Training Setups

All CFPs were preprocessed through a standardized pipeline to ensure consistency across datasets and to enhance lesion visibility for model training. Each image was resized to 224×224 pixels after removing dark borders and non-retinal background, retaining only the valid retinal region. This procedure reduced illumination artifacts from image edges and ensures spatial uniformity across samples. To highlight pathological structures such as blood vessels and microaneurysms, we employed a Ben Graham–style enhancement^[59]^ commonly used in retinal image analysis. Each image was smoothed with Gaussian blurring and combined with its original version through weighted contrast adjustment, suppressing low-frequency background and enhancing fine vascular and lesion details. During training, random horizontal and vertical flips were applied to improve robustness to spatial and acquisition variability. For the pathological tissue classification task, we strictly followed the data preprocessing pipeline employed by the UNI foundation model, specifically aligning with the protocol used for the TCGA Uniform Tumor training data. All input ROIs were resized to a spatial resolution of 224×224 pixels. Subsequently, intensity normalization was applied using standard ImageNet statistics. These augmentations mimic differences in image orientation and patient positioning, enhancing the model’s generalization across heterogeneous clinical datasets.

### Protocol Workflow

Here, we summarize the roles for each of the participating parties and their main functionalities in the FOCAL protocol as shown in Extended Data Figure 1. The protocol operates under an honest-majority threat model over the CKKS FHE scheme^[29]^. Note that the same protocol also works for other FHE schemes, such as BFV, BGV and TFHE^[65–67]^. and involves four parties: the Board of Governors (BOG), Data Contributors (DC), the AI center, and User. In what follows, we outline the main functionality for each of the participating parties.

#### Board of Governors (BOG)

A committee of *n* members responsible for the management of key generation, key management and data decryption. The BOG operates on a threshold FHE scheme, where the decryption key is distributed among its members. The keys of FOCAL are established through a distributed key generation (DKG) protocol^[68]^, a form of secure multi-party computation (MPC). This process results in *n* shares of the secret key (*sk*_1_, *sk*_2_, …, *sk*_*n*_). Note that, by having a distributed set of keys, the global private key is never reconstructed inside a single party. Consequently, any decryption operation necessitates a threshold-based decryption protocol, requiring the participation of at least 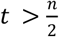 members in the BOG, where each member performs a local partial decryption using the respective share of the secret key. The main functionalities are summarized as follows.

1. Key Generation: *pk*_*g*_, (*sk*_1_, *sk*_2_, …, *sk*_*n*_) ← KeyGen (*n*). On input a parameter *n* (where *n* represents the number of members in the BOG), this algorithm outputs a global public key *pk*_*g*_ and *n* secret key share (*sk*_1_, *sk*_2_, …, *sk*_*n*_).
2. Auth-decryption: *pt*_*i*_ ← AuthDec (*ct, sk*_*i*_). On input ciphertext *ct* and a secret key share *sk*_*i*_, this algorithm outputs partial decryption share *pt*_*i*_.
3. Re-encryption: 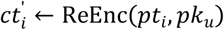. On input a decryption share *pt*_*i*_ and a user-generated public key *pk*_*u*_, this algorithm outputs a ciphertext share 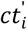 for the user.

#### Data Contributor (DC)

A data contributor (e.g., a hospital) holds a pre-trained encoder model and a local dataset contributing to the model training. In the protocol, DC processes the local private data using the pre-trained encoder model to extract a compact feature embeddings locally. Then, DC encrypts both the generated feature embedding and corresponding label using the global public key. These steps ensure that no raw data nor the feature embeddings ever leaves the DC’s premises, and that the confidentiality of the feature embeddings as well as labels can be protected during transmission and subsequent processing. The main functionalities are summarized as follows.

1. Generate Embedding: *v* ← GenEmb (*data, encoder*). On input the private datasets *data* and a pretrained encoder model *encoder*, this algorithm outputs a feature embedding vector *v*.
2. Encryption: 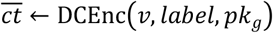. On input feature embedding vector *v*, corresponding label and the global public key *pk*_*g*_, this algorithm outputs a ciphertext tuple 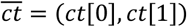 as the ciphertext, where *ct*[0] is encrypted embedding and *ct*[1] is encrypted label.

#### AI Center

The AI center is the computational server for model training and inference. The main functionality of the AI center includes a homomorphic training algorithm on the encrypted feature embedding and the corresponding label to produce a trained model entirely over ciphertexts, and an inference algorithm that performs homomorphic inference on the encrypted model and encrypted user feature embedding, producing an encrypted result. The main functionalities are summarized as follows.

1. Homomorphic Training: 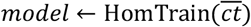. On input a part 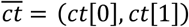, this algorithm outputs a model with a plaintext encoder and an encrypted task head.
2. Homomorphic Inference: 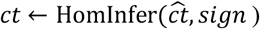. On input an encrypted embedding ciphertext 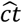 and a digital signature *sign*, this algorithm outputs encrypted inference result *ct* with a signature.

#### User

The user can either be individuals with limited computing resources or institutions with adequate computing resources. The main objective of a user is to use the trained FOCAL model. Thus, the user first generates a public-private key pair for secure communication and a digital signature for identity-checking. The user then registers the signature to the BOG for registration and whitelisting. Next, the pre-trained encoder model is run locally to generate the feature embeddings, which are then encrypted and sent to the AI center. The main functionalities are summarized as follows.

1. Register:(*pk*_*u*_, *sk*_*u*_, *sign*) ← Reg(*Identity*). On input identity of the user, this algorithm outputs a public key *pk*_*u*_, a secret key *sk*_*u*_ and a digital signature *sign*.
2. Generate Embedding: *v* ← GenEmb(*data, encoder*). On input private datasets *data* and a pre-trained encoder model *encoder*, this algorithm outputs a feature embedding vector *v*.
3. Encryption: 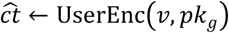. On input feature embedding vector *v* and the global public key *pk*_*g*_, this algorithm outputs an encrypted version of the feature embedding 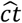.
4. Decryption: 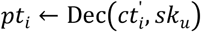. On input a label ciphertext 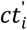 and a communication secret key share *sk*_*u*_, this algorithm outputs partial decryption share *pt*_*i*_.
5. Reconstruction: *pt* ← Reconstrut(*pt*_1_, *pt*_2_, …, *pt*_*t*_). On input *t* decryption shares (*pt*_1_, *pt*_2_, …, *pt*_*t*_), this algorithm recovers the full plaintext label *pt*.

In addition to the participating parties, the FOCAL protocol operates in two distinct phases: the training phase and the inference phase. The training phase consists of the following steps.

1. System Initialization: The BOG executes the KeyGen(*n*) to generate the global public key *pk*_*g*_ and the secret key shares (*sk*_1_, *sk*_2_, …, *sk*_*n*_). The global public key *pk*_*g*_ is published system-wide.
2. Data Preparation and Upload: Each DC executes GenEmb(*data, encoder*) to generate feature embedding vector. Next, the DC runs DCEnc(*v, label, pk*_*g*_) to output the encryption results 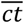 to the AI center.
3. Homomorphic Model Training: The AI Infra runs HomTrain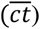 and outputs a encrypted task head.

The inference phase consists of the following steps:

1. User Registration: A user runs Reg(*dentity*) and sends *pk*_*u*_ and the signature to the BOG. The BOG registers the identity of the user to an authorized decryption list.
2. Secure Inference Request: The user uses GenEmb(*data, encoder*) to generate feature embedding vector *v*. Then UserEnc(*v, pk*_*g*_) is executed to produce the encrypted feature embedding 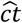. The user appends their signature to 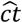, and sends the data to the AI center.
3. Homomorphic Inference: The AI center executes 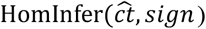 on the encrypted task head to obtain an encrypted inference result *ct* that will be forwarded to the BOG.
4. Threshold Decryption and Re-encryption: The BOG verifies the signature of the user and the signature associated with the encrypted inference result. Upon successful authentication, the BOG coordinates at least *t* members for a threshold decryption. Each member executes AuthDec(*ct, sk*_*i*_) to produce a partial decryption share *pt*_*i*_ and then uses ReEnc(*pt*_*i*_, *pk*_*u*_) to encrypt *pt*_*i*_ under the public key of the user. The encrypted partial decryption results are then sent to the user.
5. Result Reconstruction: The user decrypts each of the received shares using 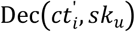 to recover the decryption share *pt*_*i*_. Finally, the user executes Reconstrut(*pt*_1_, *pt*_2_, …, *pt*_*t*_) to obtain the final plaintext inference result *pt*.

### Model Architectures

We adopted RETFound and UNI as the base models, which are built upon a ViT-Large backbone consisting of 24 transformer blocks with an embedding dimension of 1,024. When determining the splitting point, moving the split layer earlier in the ViT model increases the number of blocks within the task head, thereby enlarging the amount of trainable encrypted parameters, but slowing down both the training and the inference processes. Conversely, moving the split layer later to the end of the model reduces the number of trainable encrypted parameters in the task head, limiting the model’s adaptability when incorporating additional datasets. As shown in Extended Data Figure 2a, the external validation results on the AOD dataset confirm that choosing a mid-range split position offers the best balance between computational cost and model adaptability. Therefore, we set the split layer to 18, dividing the model into two functional components: a pretrained public encoder (blocks 1–18) and an encrypted task head (blocks 19–24 and the classification head), as illustrated in Extended Data Figure 2b. The pretrained encoder remains frozen and serves as a feature extractor, providing robust retinal representations derived from large-scale pretraining. The task head is responsible for downstream adaptation and encrypted fine-tuning. During training, only the parameters within this task head are updated using low-rank adaptation (LoRA), while the remaining layers stay fixed. This splitting strategy ensures that, during model training, the data contributors performs only feature extractions with public pretrained weights, whereas all training operations occur over ciphertexts on the AI center, thereby preserving data privacy and improving training efficiency.

### Implementation Details

We adopt the pretrained FMs, RETFound and UNI with certain modifications. The CFPs and ROIs were resized to 224×224 pixels with a patch size of 14×14 and 16×16, respectively. The AdamW optimizer was employed with a base learning rate of 5×10^-3^, cosine annealing decay, and label smoothing for regularization. Each training ran lasts for 40 epochs, with the first 10 epochs dedicated to learning rate warm-up. A batch size of 16 was used to accommodate encrypted training constraints. During training, we dynamically adjusted the classification thresholds for each category to optimize multi-class prediction performance under encrypted optimization.

### PEFT Strategy

We employed a Parameter-Efficient Fine-Tuning (PEFT) strategy^[69,70]^ to adapt the pretrained retinal FM for downstream clinical tasks under encrypted training. In this approach, the pretrained backbone remains frozen, and only a small subset of parameters within the task head are updated (approximately 0.52% of the overall model parameters), thereby minimizing the computational and communication overhead during FHE computations.

Low-Rank Adaptation (LoRA) was employed as the PEFT method due to its efficiency and compatibility with ciphertext computation. LoRA re-parameterizes the weight update of linear layers into two low-rank matrices. This decomposition captures task-specific adaptations in a compact subspace, enabling fine-tuning with only a small number of additional parameters. To maintain FHE compatibility, LoRA modules were applied only to the query (Q) and value (V) projection matrices within the self-attention layers of the task head. This design ensures that all operations remain linear, avoiding nonlinear transformations that are inefficient in encrypted computation. The LoRA rank was set to 64 with a scaling factor of 128. During training, only these LoRA parameters and the classifier were updated, while all other model weights remain fixed, which achieves efficient, privacy-preserving FM adaptation with substantially reduced number of trainable encrypted parameters.

### iDLG Attack

In conventional FL, the model weights and gradients are directly shared between the clients and the aggregating server, enabling the iDLG attack to recover sensitive samples from a single round of model update. This highlights the inherent privacy risk of gradient-based collaboration.

The iDLG attack reconstructs private input data from shared gradients. Given model parameters *θ* and a gradient ∇_*θ*_ℒ(*x*_*real*_, *y*_*real*_; *θ*) computed on private data, the attacker optimizes a dummy input *x*^’^ and label *y*^’^ to minimize

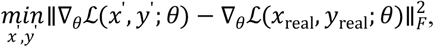

thereby reconstructing the original image.

We evaluated the vulnerability of both conventional FL and FOCAL against the iDLG attack to assess their resilience to privacy leakage, as shown in Extended Data Figure 3. The attack targeted gradients from the first block of ViT_Tiny/16, reconstructing 32×32 retinal images over 400 iterations. Here, we consider an iDLG attack to be successful if the reconstructed image has a SSIM greater than 0.5 when compared to the original input CFP. To examine the concrete privacy leakage, we performed the iDLG attack on 500 CFPs from the AOD dataset, repeating each attack run on FL and FOCAL with 150 random seeds. It is discovered that, under conventional FL, 90.6% of the reconstructed images achieved SSIMs greater than 0.5 when compared to the original input CFPs, indicating successful recoveries. Whereas, as illustrated in Figure 2f and 2g, the same iDLG method yields a 0% attack success rate, demonstrating strong resistance to gradient-based privacy leakage.

## Data Availability

All data produced in the present study are available upon reasonable request to the authors.

## References

1. Lu, M.Y., et al. A visual-language foundation model for computational pathology. Nat Med 30, 863–874 (2024). 10.1038/s41591-024-02856-4

2. Wang, X., et al. A pathology foundation model for cancer diagnosis and prognosis prediction. Nature 634, 970–978 (2024). 10.1038/s41586-024-07894-z

3. Xu, H., et al. A whole-slide foundation model for digital pathology from real-world data. Nature 630, 181–188 (2024). 10.1038/s41586-024-07441-w

4. Higgins, K., et al. The Helicobacter pylori AI-clinician harnesses artificial intelligence to personalise H. pylori treatment recommendations. Nat Commun 16, 6472 (2025). 10.1038/s41467-025-61329-5

5. Wang, M., et al. Enhancing diagnostic accuracy in rare and common fundus diseases with a knowledge-rich vision-language model. Nature Communications 16, 5528 (2025).

6. Hua, S., et al. PathoDuet: Foundation models for pathological slide analysis of H&E and IHC stains. Medical Image Analysis 97, 103289 (2024).

7. Zhou, Y., et al. A foundation model for generalizable disease detection from retinal images. Nature 622, 156–163 (2023). 10.1038/s41586-023-06555-x

8. Qiu, J., et al. Development and validation of a multimodal multitask vision foundation model for generalist ophthalmic artificial intelligence. NEJM AI 1, AIoa2300221 (2024).

9. Wu, Y., et al. An eyecare foundation model for clinical assistance: a randomized controlled trial. Nature medicine, 1–10 (2025).

10. Sun, Y., et al. A data-efficient strategy for building high-performing medical foundation models. Nature Biomedical Engineering, 1–13 (2025).

11. Tham, Y.C., et al. Building the world’s first truly global medical foundation model. Nature medicine, 1–6 (2025).

12. Luxton, D.D., et al. mHealth data security: The need for HIPAA-compliant standardization. Telemedicine and e-Health 18, 284–288 (2012).

13. Protection, F.D. General data protection regulation (GDPR). Intersoft Consulting, Accessed in October 24 (2018).

14. Yang, Y., et al. A digital mask to safeguard patient privacy. Nature medicine 28, 1883–1892 (2022).

15. Ting, D.S.W., et al. Artificial intelligence and deep learning in ophthalmology. British Journal of Ophthalmology 103, 167–175 (2019).

16. Moor, M., et al. Foundation models for generalist medical artificial intelligence. Nature 616, 259–265 (2023). 10.1038/s41586-023-05881-4

17. Lin, D., et al. Application of Comprehensive Artificial intelligence Retinal Expert (CARE) system: a national real-world evidence study. The Lancet Digital Health 3, e486–e495 (2021).

18. Kermany, D.S., et al. Identifying medical diagnoses and treatable diseases by image-based deep learning. cell 172, 1122–1131. e9 (2018).

19. Ursin, G., et al. Sharing data safely while preserving privacy. Lancet 394, 1902 (2019). 10.1016/S0140-6736(19)32603-0

20. Kaissis, G.A., et al. Secure, privacy-preserving and federated machine learning in medical imaging. Nature Machine Intelligence 2, 305–311 (2020). 10.1038/s42256-020-0186-1

21. Pati, S., et al. Federated learning enables big data for rare cancer boundary detection. Nature communications 13, 7346 (2022).

22. Kaissis, G., et al. End-to-end privacy preserving deep learning on multi-institutional medical imaging. Nature Machine Intelligence 3, 473–484 (2021).

23. Li, T., et al. Federated learning: Challenges, methods, and future directions. IEEE signal processing magazine 37, 50–60 (2020).

24. Zhu, L., et al. Deep leakage from gradients. Advances in neural information processing systems 32 (2019).

25. Zhao, B., et al. iDLG: Improved deep leakage from gradients. arXiv preprint arXiv:2001.02610 (2020).

26. Hatamizadeh, A., et al. Gradvit: Gradient inversion of vision transformers. in Proceedings of the IEEE/CVF conference on computer vision and pattern recognition. 10021–10030.

27. Imteaj, A., et al. A survey on federated learning for resource-constrained IoT devices. IEEE Internet of Things Journal 9, 1–24 (2021).

28. Bian, S., et al. HE3DB: An efficient and elastic encrypted database via arithmetic-and-logic fully homomorphic encryption. in Proceedings of the 2023 ACM SIGSAC Conference on Computer and Communications Security. 2930–2944.

29. Cheon, J.H., et al. Homomorphic encryption for arithmetic of approximate numbers. in International conference on the theory and application of cryptology and information security. 409–437 (Springer).

30. Thapa, C., et al. Splitfed: When federated learning meets split learning. In Proceedings of the AAAI conference on artificial intelligence. 8485–8493.

31. Dwork, C. Differential privacy. in International colloquium on automata, languages, and programming. 1–12 (Springer).

32. Liu, J., et al. Recoverable privacy-preserving image classification through noise-like adversarial examples. ACM Transactions on Multimedia Computing, Communications and Applications 20, 1–27 (2024).

33. Li, J., et al. Privacy-preserving portrait matting. in Proceedings of the 29th ACM international conference on multimedia. 3501–3509.

34. Goldreich, O. Foundations of Cryptography, Volume 2. (Cambridge university press Cambridge, 2004).

35. Micali, S.G.a.S. Probabilistic encryption. Journal of Computer and System Sciences 28, 270–299 (1984).

36. Jauernig, P., et al. Trusted execution environments: properties, applications, and challenges. IEEE Security & Privacy 18, 56–60 (2020).

37. Hu, E.J., et al. Lora: Low-rank adaptation of large language models. ICLR 1, 3 (2022).

38. Wang, H., et al. Score-CAM: Score-weighted visual explanations for convolutional neural networks. in Proceedings of the IEEE/CVF conference on computer vision and pattern recognition workshops. 24–25.

39. Vaswani, A., et al. Attention is all you need. Advances in neural information processing systems 30 (2017).

40. Kaggle. Diabetic Retinopathy Detection Challenge, <https://www.kaggle.com/datasets/nurmukhammed7/augemnted-ocular-diseases> (2015).

41. Pachade, S., et al. Retinal fundus multi-disease image dataset (rfmid): A dataset for multi-disease detection research. Data 6, 14 (2021).

42. Ting, D.S.W., et al. Development and validation of a deep learning system for diabetic retinopathy and related eye diseases using retinal images from multiethnic populations with diabetes. Jama 318, 2211–2223 (2017).

43. Ong, K.L., et al. Global, regional, and national burden of diabetes from 1990 to 2021, with projections of prevalence to 2050: a systematic analysis for the Global Burden of Disease Study 2021. The Lancet 402, 203–234 (2023).

44. Dai, L., et al. A deep learning system for predicting time to progression of diabetic retinopathy. Nature Medicine 30, 584–594 (2024).

45. Lin, L., et al. The SUSTech-SYSU dataset for automated exudate detection and diabetic retinopathy grading. Scientific Data 7, 409 (2020).

46. Platform, A.T. Diabetic Retinopathy Arranged Dataset, <https://tianchi.aliyun.com/dataset/93926> (2023).

47. Chen, R.J., et al. Towards a general-purpose foundation model for computational pathology. Nature medicine 30, 850–862 (2024).

48. Tolkach, Y., et al. Artificial intelligence for tumour tissue detection and histological regression grading in oesophageal adenocarcinomas: a retrospective algorithm development and validation study. The Lancet Digital Health 5, e265–e275 (2023).

49. Holmberg, O.G., et al. Self-supervised retinal thickness prediction enables deep learning from unlabelled data to boost classification of diabetic retinopathy. Nature Machine Intelligence 2, 719–726 (2020).

50. Engelmann, J., et al. Detecting multiple retinal diseases in ultra-widefield fundus imaging and data-driven identification of informative regions with deep learning. Nature Machine Intelligence 4, 1143–1154 (2022).

51. Kaggle. Cataract Dataset, <https://www.kaggle.com/datasets/jr2ngb/cataractdataset> (2020).

52. Nakayama, L.F., et al. A Brazilian multilabel ophthalmological dataset (BRSET). PhysioNet 13026 (2023).

53. Benítez, V.E.C., et al. Dataset from fundus images for the study of diabetic retinopathy. Data in brief 36, 107068 (2021).

54. Orlando, J.I., et al. Refuge challenge: A unified framework for evaluating automated methods for glaucoma assessment from fundus photographs. Medical image analysis 59, 101570 (2020).

55. Liu, R., et al. Deepdrid: Diabetic retinopathy—grading and image quality estimation challenge. Patterns 3 (2022).

56. Kumar, J.H., et al. Chákşu: A glaucoma specific fundus image database. Scientific data 10, 70 (2023).

57. Zhang, Z., et al. Origa-light: An online retinal fundus image database for glaucoma analysis and research. in 2010 Annual international conference of the IEEE engineering in medicine and biology. 3065–3068 (IEEE).

58. Porwal, P., et al. Idrid: Diabetic retinopathy–segmentation and grading challenge. Medical image analysis 59, 101561 (2020).

59. Chatpatanasiri, R. APTOS: Eye Preprocessing in Diabetic Retinopathy, <https://www.kaggle.com/ratthachat/aptos-eye-preprocessing-in-diabetic-retinopathy> (2019).

60. Jin, K., et al. Fives: A fundus image dataset for artificial intelligence based vessel segmentation. Scientific data 9, 475 (2022).

61. Kovalyk, O., et al. PAPILA: Dataset with fundus images and clinical data of both eyes of the same patient for glaucoma assessment. Scientific Data 9, 291 (2022).

62. Diaz-Pinto, A., et al. CNNs for automatic glaucoma assessment using fundus images: an extensive validation. Biomedical engineering online 18, 29 (2019).

63. Decencière, E., et al. Feedback on a publicly distributed image database: the Messidor database. Image Analysis & Stereology, 231–234 (2014).

64. Bajwa, M.N., et al. G1020: A benchmark retinal fundus image dataset for computer-aided glaucoma detection. in 2020 International Joint Conference on Neural Networks (IJCNN). 1–7 (IEEE).

65. Brakerski, Z., et al. (Leveled) fully homomorphic encryption without bootstrapping. ACM Transactions on Computation Theory (TOCT) 6, 1–36 (2014).

66. Brakerski, Z. Fully homomorphic encryption without modulus switching from classical GapSVP. in Annual cryptology conference. 868–886 (Springer).

67. Chillotti, I., et al. TFHE: fast fully homomorphic encryption over the torus. Journal of Cryptology 33, 34–91 (2020).

68. Gennaro, R., et al. Secure Distributed Key Generation for Discrete-Log Based Cryptosystems. Journal of Cryptology 20, 51–83 (2006). 10.1007/s00145-006-0347-3

69. Liu, H., et al. Few-shot parameter-efficient fine-tuning is better and cheaper than in-context learning. Advances in Neural Information Processing Systems 35, 1950–1965 (2022).

70. Ding, N., et al. Parameter-efficient fine-tuning of large-scale pre-trained language models. Nature machine intelligence 5, 220–235 (2023).

